# Rapid emergence of UV stabilizer Bis(2,2,6,6-tetramethyl-4-piperidyl) sebacate (BTMPS) in the illicit fentanyl supply across the United States in July-August 2024: Results from drug and drug paraphernalia testing

**DOI:** 10.1101/2024.09.13.24313643

**Authors:** Chelsea L. Shover, Morgan E. Godvin, Meghan Appley, Elise M. Pyfrom, Fernando Montero Castrillo, Karli Hochstatter, Talia Nadel, Neil Garg, Adam Koncsol, Joseph R. Friedman, Caitlin A. Molina, Ruby Romero, Brendan Harshberger, Jordan Spoliansky, Sarah Laurel, Elham Jalayer, Juan Ruelas, John Gonzales, Soma Snakeoil, Sonya Guerra, Oscar Arellano, Candace Winstead, Margaret Rybak, Joanna Champney, Brent Waninger, Edward Sisco

**Affiliations:** Division of General Internal Medicine and Health Services Research, University of California, Los Angeles, CA, USA; National Institute of Standards and Technology, Gaithersburg, MD, USA; Department of Anthropology, Columbia University, New York, NY, USA; Friends Research Institute, Baltimore, MD, USA; Chemistry and Biochemistry, University of California, Los Angeles, CA, USA; Department of Psychiatry, University of California, San Diego, CA, USA; Los Angeles County Department of Health Services, Los Angeles, CA, USA; Savage Sisters Recovery, Philadelphia, PA, USA; Bienestar, Los Angeles, CA, USA; The Sidewalk Project, Los Angeles, CA, USA; Homeless Outreach Program’s Integrated Care System (HOPICS), Los Angeles, CA, USA; California State Polytechnic University, San Luis Obispo, CA, USA; Center for Harm Reduction Services, Maryland Department of Health, Baltimore, MD, USA; Delaware Division of Substance Abuse and Mental Health, New Castle, DE, USA

## Abstract

**Background:** Changes to the US drug supply historically unfold slowly with predictable patterns of geographic diffusion. Here we draw on drug checking results from around the United States to report a rapid shift in the illicit drug supply with important implications for public health. Bis(2,2,6,6-tetramethyl-4-piperidyl) sebacate, or “BTMPS” is a hindered amine light stabilizer with various industrial applications. Animal studies indicate multiple kinds of adverse health effects.

**Methods:** Drug samples collected by community-based drug checking programs in Los Angeles and Philadelphia, along with drug residue samples from other jurisdictions in California, Delaware, Maryland, and Washington, were submitted to the National Institute on Standards and Technology. Samples were qualitatively tested with Direct Analysis in Real Time mass spectrometry, with reflex to liquid chromatography mass spectrometry (LCMS) for confirmatory testing. Quantitation – percent by mass – was performed using LCMS. At both sites where drug samples were collected, participants were asked to respond to a survey that included questions about what the substance was sold as and any unexpected effects.

**Results:** Between June 1, 2024 and August 31, 2024, a total of 178 samples sold as fentanyl were tested. Of these, 43 (24%) contained BTMPS, with the proportion per month rising from 0% in June to more than a third in August. An additional 23 residue samples from sites doing residue testing contained BTMPS. Fentanyl samples with BTMPS also contained many other compounds, including local anesthetics and alpha-2 agonists. Average fentanyl purity was significantly lower in samples with BTMPS compared to samples without.

**Conclusions:** The introduction of an industrial chemical to the illicit drug supply at this speed and scale is unprecedented and concerning. Further research is urgently needed to determine why it is present in the fentanyl supply and characterize effects on human health.

## Introduction

Changes to the U.S. illicit drug supply historically unfold slowly, with predictable patterns of geographic diffusion.^1, 2^ Illicitly manufactured fentanyl was introduced into the heroin supply on the east coast and Appalachia in the early 2010’s, but took about a decade to become a major part of the illicit drug supply in the western U.S.^2^ Xylazine was first added to heroin in Puerto Rico, then emerged in Philadelphia as an additive to heroin with fentanyl following, before spreading to jurisdictions across the U.S. over a decade.^1^ Here we report evidence of a more sudden change in the illicit drug supply and examine the potential health and market implications.

Bis(2,2,6,6-tetramethyl-4-piperidyl) sebacate, or “BTMPS” is a hindered amine light stabilizer used in plastics manufacturing, as an adhesive, and as a sealant.^3-6^ Animal studies have revealed adverse health effects upon exposure to BTMPS at relatively high doses, including cardiotoxicity and sudden death.^7^ BTMPS has been found to be pharmacologically active as a nicotinic antagonist in animal studies; however this compound has not been approved for human consumption, nor has administration to humans been studied.^3-5^

Drawing on a multisite drug testing program, we characterize this industrial chemical’s introduction to the illicit drug supply, almost simultaneously, in multiple locations around the U.S.

## Methods

The samples discussed here were obtained from community-based drug checking programs. Through these programs, a small amount (trace residue to a few milligrams) of an individual’s drugs are collected and tested – allowing for collection of samples that provide insight into the illicit drug supply not available through other data sources.^8^ Two sample collection approaches were used. For trace residue sample collection, cotton swabs or meta-aramid wipes were used to wipe either a piece of used paraphernalia (syringe, cooker, baggie, etc.) or actual drug product to collect residual particulate for analysis. For drug product collection, a few milligrams of actual drug product (*i.e*., powder or crushed pill) were collected using a microscoop and transferred to a 2 mL vial containing acetonitrile. Collected samples were provided unique identifiers and were submitted to the National Institute of Standards and Technology (NIST) for analysis. All samples were qualitatively tested with direct analysis in real time mass spectrometry (DART-MS) and the resulting spectra searched against a library of over 1,300 compounds including drugs, cutting agents, and adulterants using previously published protocols.^9, 10^ Quantitative analysis was completed on all drug product samples using liquid chromatography tandem mass spectrometry (LC-MS/MS) with results reported back as percent by mass. The quantitation panel included compounds across several classes: fentanyl and fentanyl analogs (fentanyl, *para*-fluorofentanyl), other common illicit drugs (cocaine, heroin, methamphetamine), fentanyl precursors (4-ANPP, phenethyl 4-ANPP), α_2_-agonists (medetomidine, xylazine), and adulterants (BTMPS, lidocaine, tetracaine). Additional information on sample preparation and instrumental analysis for quantitation can be found in the **Supplemental Information**. For statistical testing using quantitative results, we inputted 0.1 % for values below the limit of quantitation.

Drug product samples were provided by community-based drug checking sites in Los Angeles, CA and Philadelphia, PA. Trace residue samples were provided by community-based drug checking sites within California (Los Angeles and other cities), Delaware, Maryland, Nevada, and Washington.

At the Los Angeles and Philadelphia sites, participants were administered a survey that included questions about what the substance was sold as, and if there were any unexpected effects. UCLA IRB determined that the activities described in this analysis, funded by CDC OD2A:LOCAL were public health surveillance and did not constitute human subjects research. The research activities in Philadelphia were approved by the WIRB-Copernicus Group (WCG IRB). No identifiable information or survey data was provided to NIST scientists.

### Statistical methods

Descriptive statistics were calculated, including percentage of fentanyl samples per month and per site containing BTMPS, and prevalence of co-detected compounds. Chi-square tests were performed to compare the prevalence of other detected compounds by presence of BTMPS, overall and stratified by site (alpha level 0.05). Among quantitated samples, mean fentanyl purity and concentration of other compounds were compared by presence of BTMPS using a two-sample t-test (alpha level 0.05).

## Results

Between June 1, 2024, and August 31, 2024, a total of 178 drug product samples sold as fentanyl were tested from Los Angeles and Philadelphia. Of these, 43 (25 %) contained BTMPS, with the proportion per month rising from 0 % in June to 36 % in August (**Figure 1**). Of the 178 samples sold as fentanyl, 175 (98 %) contained fentanyl. One contained only lidocaine (3.3 % by weight), and two contained fentanyl analogs or precursors but not fentanyl. BTMPS was not identified in any sample prior to June 2024.

**Figure 1.**
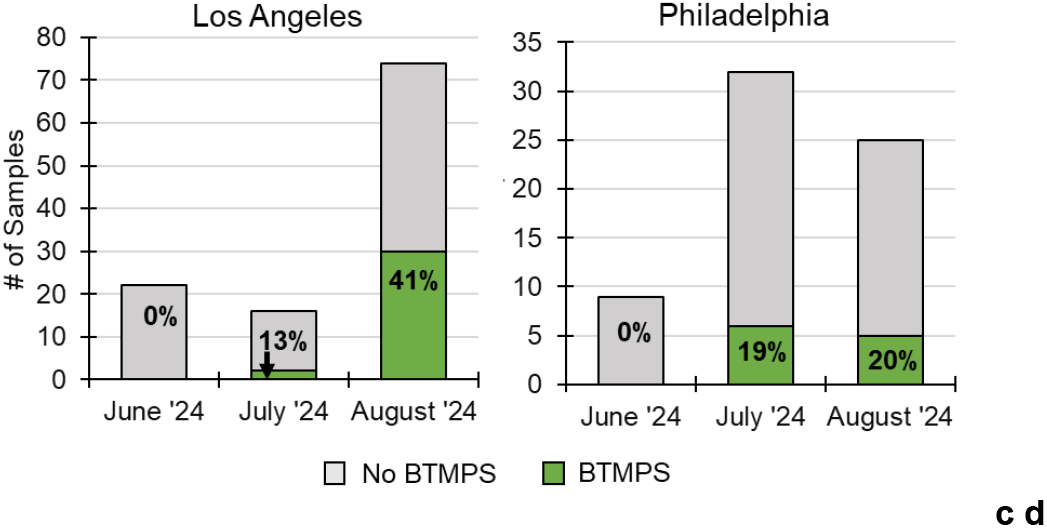
Number and percentage of tested samples sold as fentanyl that contained BTMPS in Los Angeles (left) and Philadelphia (right) between June 2024 and August 2024.

Fentanyl samples with BTMPS also contained other compounds, with substantial heterogeneity between samples and site (**Figure 2, Table 1, and Table 2**). Alpha-2 agonists were present in fentanyl samples from both sites, with xylazine significantly more likely to be present in samples with BTMPS compared to fentanyl without BTMPS. Eight of the 11 Philadelphia BTMPS samples also contained medetomidine, which was not detected in Los Angeles. In the quantitative results, xylazine concentration was not significantly correlated with BTMPS presence. Fluorofentanyl was significantly more common in BTMPS samples, particularly in Los Angeles, where half of the BTMPS samples also contained this fentanyl analog. 4-ANPP, a fentanyl precursor used in several of the most common synthesis methods was present in 84 % of samples and was particularly ubiquitous in Los Angeles at 94 % (n=30), compared to 55 % (n=6) in Philadelphia. Two other fentanyl precursor chemicals, N-phenethyl-N-phenylpropionamide and bipiperidinyl 4-ANPP, were only found in samples that contained BTMPS.

**Table 1.** Characteristics of samples of drugs sold as fentanyl, stratified by presence of BTMPS.

| | Contains<br>BTMPS | | Does not contain<br>BTMPS | | $\chi^2$ | Total | |
| --- | --- | --- | --- | --- | --- | --- | --- |
|  | *n* | % | *n* | % | *p* | *n* | % |
| **Drug checking site** |  |  |  |  | **0.1** |  |  |
| Philadelphia | 11 | 26% | 55 | 41% |  | 66 | 37% |
| Los Angeles | 32 | 74% | 80 | 59% |  | 112 | 63% |
| **Detected substances** |  |  |  |  |  |  |  |
| Bis(2,2,6,6-tetramethyl-4-piperidyl)sebacate | 43 | 100% | 0 | 0% | N/A | 43 | 24% |
| **Fentanyl and fentanyl analogs** |  |  |  |  |  |  |  |
| Fentanyl | 43 | 100% | 132 | 98% | 1 | 175 | 98% |
| Fluorofentanyl | 16 | 37% | 26 | 19% | 0.02 | 42 | 23% |
| Methylfentanyl | 0 | 0% | 2 | 2% | 0.4 | 2 | 1% |
| **Fentanyl and fentanyl analog precursors** |  |  |  |  |  |  |  |
| 4-ANPP | 36 | 84% | 96 | 71% | 0.1 | 132 | 74% |
| Phenethyl 4-ANPP | 16 | 37% | 32 | 24% | 0.1 | 48 | 27% |
| Aniline | 9 | 21% | 18 | 13% | 0.3 | 27 | 15% |
| Despropionyl Fluorofentanyl | 8 | 19% | 14 | 10% | 0.2 | 22 | 12% |
| Norfentanyl | 5 | 12% | 15 | 11% | 1 | 20 | 11% |
| N-Phenethyl-N-phenylpropionamide | 4 | 9% | 0 | 0% | <0.001 | 4 | 2% |
| Ethyl 4-ANPP | 2 | 5% | 14 | 10% | 0.2 | 16 | 9% |
| Bipiperidinyl 4-ANPP | 1 | 2% | 0 | 0% | 0.1 | 1 | 1% |
| NPP | 0 | 0% | 4 | 3% | 0.2 | 4 | 2% |
| 4-Anilino-1-boc-piperidine | 0 | 0% | 3 | 2% | 0.3 | 3 | 2% |
| 1-Phenethylpiperidin-4-ol | 0 | 0% | 2 | 1% | 0.4 | 2 | 1% |
| Despropionyl Methylfentanyl | 0 | 0% | 2 | 1% | 0.4 | 2 | 1% |
| **$\alpha_2$-Agonists** |  |  |  |  |  |  |  |
| Xylazine | 22 | 51% | 39 | 29% | 0.01 | 61 | 34% |
| Medetomidine | 8 | 19% | 32 | 24% | 0.5 | 40 | 22% |
| Etomidate | 0 | 0% | 1 | 1% | 0.6 | 1 | 1% |
| **Non-fentanyl opioids** |  |  |  |  |  |  |  |
| Tramadol | 2 | 5% | 4 | 3% | 0.6 | 6 | 3% |
| Heroin | 0 | 0% | 3 | 2% | 1 | 3 | 2% |
| N-desethyl Etonitazene | 0 | 0% | 1 | 1% | 0.6 | 1 | 1% |
| **Cocaine and local anesthetics** |  |  |  |  |  |  |  |
| Cocaine | 2 | 5% | 5 | 4% | 0.8 | 7 | 4% |
| Benzoylcegonine | 1 | 2% | 0 | 0% | 0.1 | 1 | 1% |
| Lidocaine | 36 | 84% | 66 | 49% | <0.001 | 102 | 57% |
| Tetracaine | 9 | 21% | 27 | 20% | 0.9 | 36 | 20% |
| Procaine | 8 | 19% | 16 | 12% | 0.3 | 24 | 13% |
| Benzocaine | 1 | 2% | 0 | 0% | 0.1 | 1 | 1% |
| Bupivacaine | 0 | 0% | 1 | 1% | 1 | 1 | 1% |
| **Other drugs** |  |  |  |  |  |  |  |
| Methamphetamine | 6 | 14% | 4 | 3% | <0.01 | 10 | 6% |
| Ketamine | 0 | 0% | 1 | 1% | 0.6 | 1 | 1% |
| Nicotine | 0 | 0% | 2 | 1% | 0.4 | 2 | 1% |
| **Cutting agents and diluents** |  |  |  |  |  |  |  |
| Mannitol | 35 | 81% | 70 | 52% | <0.001 | 105 | 59% |
| Acetaminophen | 24 | 56% | 47 | 35% | 0.01 | 71 | 40% |
| Caffeine | 16 | 37% | 32 | 24% | 0.1 | 48 | 27% |
| Phenacetin | 2 | 5% | 1 | 1% | 0.1 | 3 | 2% |
| Diphenhydramine | 1 | 2% | 0 | 0% | 0.1 | 1 | 1% |
| Theobromine | 0 | 0% | 1 | 1% | 0.6 | 1 | 1% |
| Nicotinamide | 0 | 0% | 2 | 1% | 0.4 | 2 | 1% |
| Metamizole | 0 | 0% | 2 | 1% | 0.4 | 2 | 1% |
| Total | 43 |  | 135 |  |  | 178 |  |

**Table 2.**
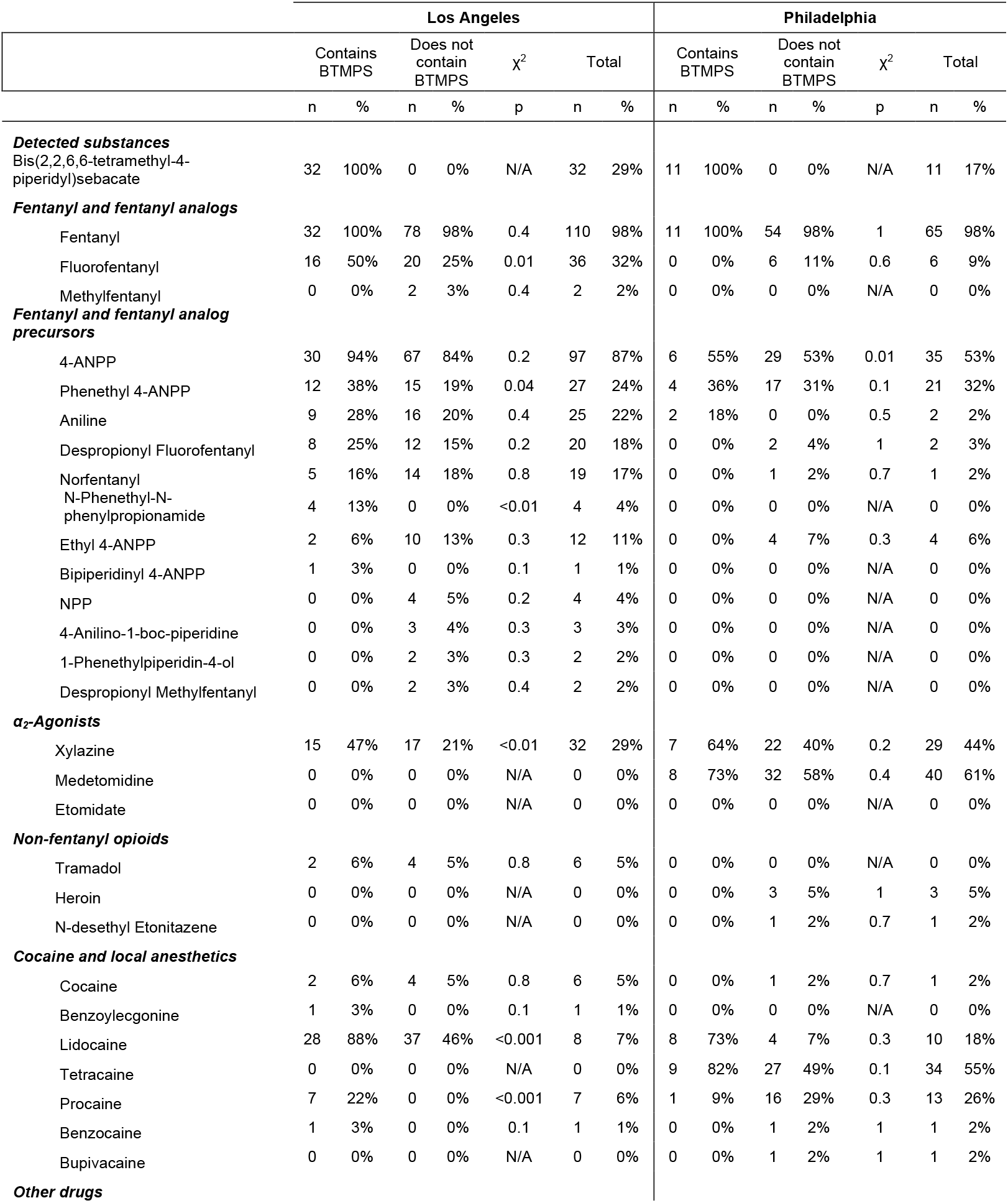
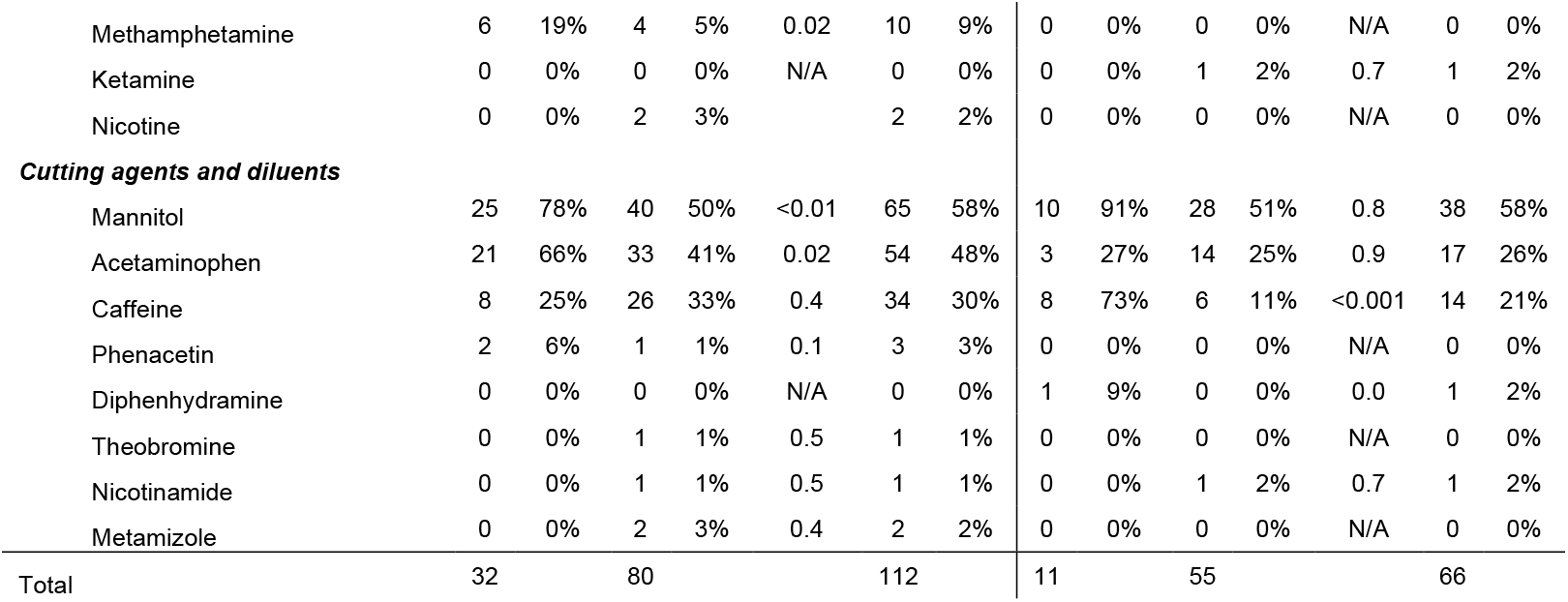
Characteristics of samples of drugs sold as fentanyl, stratified by presence of BTMPS and location.

| Los Angeles |  |  |  |  |  |  |  |  | Philadelphia |  |  |  |  |  |  |
| --- | --- | --- | --- | --- | --- | --- | --- | --- | --- | --- | --- | --- | --- | --- | --- |
| | Contains BTMPS | | Does not contain BTMPS | | $\chi^2$ | Total | | | Contains BTMPS | | Does not contain BTMPS | | $\chi^2$ | Total | |
|  | n | % | n | % | p | n | % |  | n | % | n | % | p | n | % |
| **Detected substances** |  |  |  |  |  |  |  |  |  |  |  |  |  |  |  |
| Bis(2,2,6,6-tetramethyl-4-piperidyl)sebacate | 32 | 100% | 0 | 0% | N/A | 32 | 29% |  | 11 | 100% | 0 | 0% | N/A | 11 | 17% |
| **Fentanyl and fentanyl analogs** |  |  |  |  |  |  |  |  |  |  |  |  |  |  |  |
| Fentanyl | 32 | 100% | 78 | 98% | 0.4 | 110 | 98% |  | 11 | 100% | 54 | 98% | 1 | 65 | 98% |
| Fluorofentanyl | 16 | 50% | 20 | 25% | 0.01 | 36 | 32% |  | 0 | 0% | 6 | 11% | 0.6 | 6 | 9% |
| Methylfentanyl | 0 | 0% | 2 | 3% | 0.4 | 2 | 2% |  | 0 | 0% | 0 | 0% | N/A | 0 | 0% |
| **Fentanyl and fentanyl analog precursors** |  |  |  |  |  |  |  |  |  |  |  |  |  |  |  |
| 4-ANPP | 30 | 94% | 67 | 84% | 0.2 | 97 | 87% |  | 6 | 55% | 29 | 53% | 0.01 | 35 | 53% |
| Phenethyl 4-ANPP | 12 | 38% | 15 | 19% | 0.04 | 27 | 24% |  | 4 | 36% | 17 | 31% | 0.1 | 21 | 32% |
| Aniline | 9 | 28% | 16 | 20% | 0.4 | 25 | 22% |  | 2 | 18% | 0 | 0% | 0.5 | 2 | 2% |
| Despropionyl Fluorofentanyl | 8 | 25% | 12 | 15% | 0.2 | 20 | 18% |  | 0 | 0% | 2 | 4% | 1 | 2 | 3% |
| Norfentanyl | 5 | 16% | 14 | 18% | 0.8 | 19 | 17% |  | 0 | 0% | 1 | 2% | 0.7 | 1 | 2% |
| N-Phenethyl-N-phenylpropionamide | 4 | 13% | 0 | 0% | <0.01 | 4 | 4% |  | 0 | 0% | 0 | 0% | N/A | 0 | 0% |
| Ethyl 4-ANPP | 2 | 6% | 10 | 13% | 0.3 | 12 | 11% |  | 0 | 0% | 4 | 7% | 0.3 | 4 | 6% |
| Bipiperidiny 4-ANPP | 1 | 3% | 0 | 0% | 0.1 | 1 | 1% |  | 0 | 0% | 0 | 0% | N/A | 0 | 0% |
| NPP | 0 | 0% | 4 | 5% | 0.2 | 4 | 4% |  | 0 | 0% | 0 | 0% | N/A | 0 | 0% |
| 4-Anilino-1-boc-piperidine | 0 | 0% | 3 | 4% | 0.3 | 3 | 3% |  | 0 | 0% | 0 | 0% | N/A | 0 | 0% |
| 1-Phenethylpiperidin-4-ol | 0 | 0% | 2 | 3% | 0.3 | 2 | 2% |  | 0 | 0% | 0 | 0% | N/A | 0 | 0% |
| Despropionyl Methylfentanyl | 0 | 0% | 2 | 3% | 0.4 | 2 | 2% |  | 0 | 0% | 0 | 0% | N/A | 0 | 0% |
| **$\alpha_2$-Agonists** |  |  |  |  |  |  |  |  |  |  |  |  |  |  |  |
| Xylazine | 15 | 47% | 17 | 21% | <0.01 | 32 | 29% |  | 7 | 64% | 22 | 40% | 0.2 | 29 | 44% |
| Medetomidine | 0 | 0% | 0 | 0% | N/A | 0 | 0% |  | 8 | 73% | 32 | 58% | 0.4 | 40 | 61% |
| Etomidate | 0 | 0% | 0 | 0% | N/A | 0 | 0% |  | 0 | 0% | 0 | 0% | N/A | 0 | 0% |
| **Non-fentanyl opioids** |  |  |  |  |  |  |  |  |  |  |  |  |  |  |  |
| Tramadol | 2 | 6% | 4 | 5% | 0.8 | 6 | 5% |  | 0 | 0% | 0 | 0% | N/A | 0 | 0% |
| Heroin | 0 | 0% | 0 | 0% | N/A | 0 | 0% |  | 0 | 0% | 3 | 5% | 1 | 3 | 5% |
| N-desethyl Etonitazene | 0 | 0% | 0 | 0% | N/A | 0 | 0% |  | 0 | 0% | 1 | 2% | 0.7 | 1 | 2% |
| **Cocaine and local anesthetics** |  |  |  |  |  |  |  |  |  |  |  |  |  |  |  |
| Cocaine | 2 | 6% | 4 | 5% | 0.8 | 6 | 5% |  | 0 | 0% | 1 | 2% | 0.7 | 1 | 2% |
| Benzoylcegonine | 1 | 3% | 0 | 0% | 0.1 | 1 | 1% |  | 0 | 0% | 0 | 0% | N/A | 0 | 0% |
| Lidocaine | 28 | 88% | 37 | 46% | <0.001 | 8 | 7% |  | 8 | 73% | 4 | 7% | 0.3 | 10 | 18% |
| Tetracaine | 0 | 0% | 0 | 0% | N/A | 0 | 0% |  | 9 | 82% | 27 | 49% | 0.1 | 34 | 55% |
| Procaine | 7 | 22% | 0 | 0% | <0.001 | 7 | 6% |  | 1 | 9% | 16 | 29% | 0.3 | 13 | 26% |
| Benzocaine | 1 | 3% | 0 | 0% | 0.1 | 1 | 1% |  | 0 | 0% | 1 | 2% | 1 | 1 | 2% |
| Bupivacaine | 0 | 0% | 0 | 0% | N/A | 0 | 0% |  | 0 | 0% | 1 | 2% | 1 | 1 | 2% |
| **Other drugs** |  |  |  |  |  |  |  |  |  |  |  |  |  |  |  |

|  |  |  |  |  |  |  |  |  |  |  |  |  |  |  |
| --- | --- | --- | --- | --- | --- | --- | --- | --- | --- | --- | --- | --- | --- | --- |
| Methamphetamine | 6 | 19% | 4 | 5% | 0.02 | 10 | 9% | 0 | 0% | 0 | 0% | N/A | 0 | 0% |
| Ketamine | 0 | 0% | 0 | 0% | N/A | 0 | 0% | 0 | 0% | 1 | 2% | 0.7 | 1 | 2% |
| Nicotine | 0 | 0% | 2 | 3% |  | 2 | 2% | 0 | 0% | 0 | 0% | N/A | 0 | 0% |
| **Cutting agents and diluents** |  |  |  |  |  |  |  |  |  |  |  |  |  |  |
| Mannitol | 25 | 78% | 40 | 50% | <0.01 | 65 | 58% | 10 | 91% | 28 | 51% | 0.8 | 38 | 58% |
| Acetaminophen | 21 | 66% | 33 | 41% | 0.02 | 54 | 48% | 3 | 27% | 14 | 25% | 0.9 | 17 | 26% |
| Caffeine | 8 | 25% | 26 | 33% | 0.4 | 34 | 30% | 8 | 73% | 6 | 11% | <0.001 | 14 | 21% |
| Phenacetin | 2 | 6% | 1 | 1% | 0.1 | 3 | 3% | 0 | 0% | 0 | 0% | N/A | 0 | 0% |
| Diphenhydramine | 0 | 0% | 0 | 0% | N/A | 0 | 0% | 1 | 9% | 0 | 0% | 0.0 | 1 | 2% |
| Theobromine | 0 | 0% | 1 | 1% | 0.5 | 1 | 1% | 0 | 0% | 0 | 0% | N/A | 0 | 0% |
| Nicotinamide | 0 | 0% | 1 | 1% | 0.5 | 1 | 1% | 0 | 0% | 1 | 2% | 0.7 | 1 | 2% |
| Metamizole | 0 | 0% | 2 | 3% | 0.4 | 2 | 2% | 0 | 0% | 0 | 0% | N/A | 0 | 0% |
| Total | 32 |  | 80 |  |  | 112 |  | 11 |  | 55 |  |  | 66 |  |

**Figure 2.**
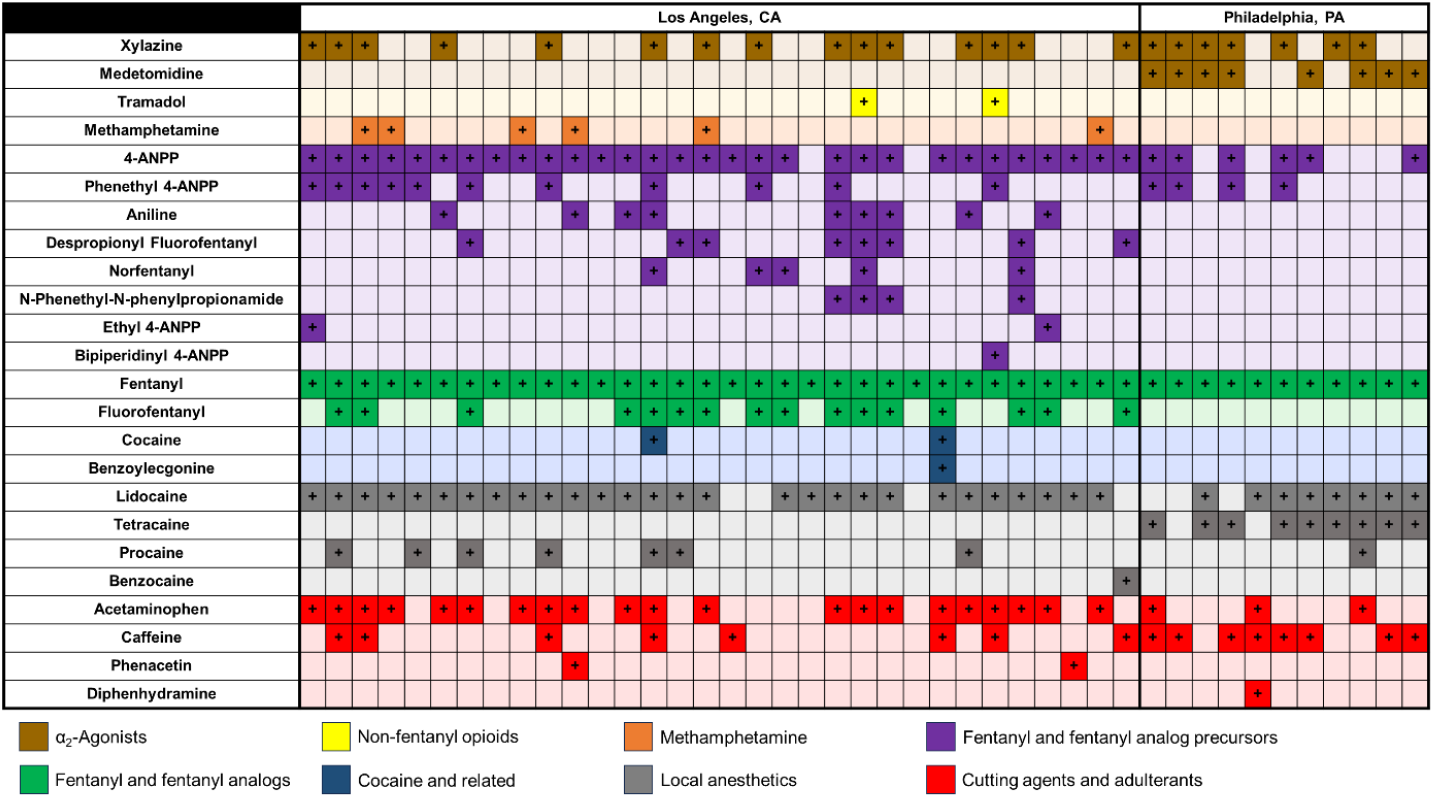
Co-detected compounds in samples that contained BTMPS. A dark shaded cell with a “+” indicates that compound was co-detected with BTMPS. Each column represents a unique sample.

Cocaine and structurally similar local anesthetics (*i.e*., lidocaine, procaine, and tetracaine) were found to be commonly co-detected in BTMPS samples. Lidocaine was prevalent in 36 (84 %) of BTMPS samples across both sites. In Los Angeles, seven (22 %) samples had procaine and two (6 %) had cocaine itself. In Philadelphia, no samples had procaine or cocaine but nine (82 %) were found to contain tetracaine. Only four (10 %) of the 43 samples containing BTMPS had no cocaine or structurally similar local anesthetic present.

### Quantitative testing

Quantitative results were available for 75 samples from Los Angeles (37 samples were trace residues) (**Figure 3**) and all 66 samples from Philadelphia. In Los Angeles, 28 (38 %) of samples contained BTMPS, with 19 detectable above the limit of quantitation (approximately 0.1 % by weight in a powder or crushed pill). BTMPS accounted for 0.8 % to 35 % by mass across the 19 samples. Mean fentanyl purity was significantly lower in samples that contained BTMPS (3.9 % by mass, 95 % CI 2.4, 5.4) compared to samples without BTMPS (9.6 % by mass, 95 % CI 5.5, 13.6). Fentanyl purity in BTMPS samples was relatively uniform and low, with only two samples exceeding 10 % by mass. In contrast, there was substantial heterogeneity in fentanyl purity of samples without BTMPS, with a range of 0 % (the sample that contained only lidocaine) to 48 % by mass. Mean 4-ANPP concentration was also significantly lower in BTMPS samples with a mean percent by mass of 0.9 %, 95 % CI 0.4, 1.3, compared to 2.6 %, 95 % CI 1.5, 3.8 in fentanyl samples without BTMPS. In Philadelphia, 11 (16 %) of quantified samples contained BTMPS, with eight having BTMPS detectable above the limit of quantitation and percent by mass ranging from 1 % to 18 %. As in Los Angeles, mean fentanyl purity in Philadelphia was significantly lower in samples with BTMPS (3.0 % by mass, 95 % CI 1.0, 4.4) compared to those without BTMPS (7.6 % by mass, 95 % CI 5.6, 9.5).

**Figure 3.**
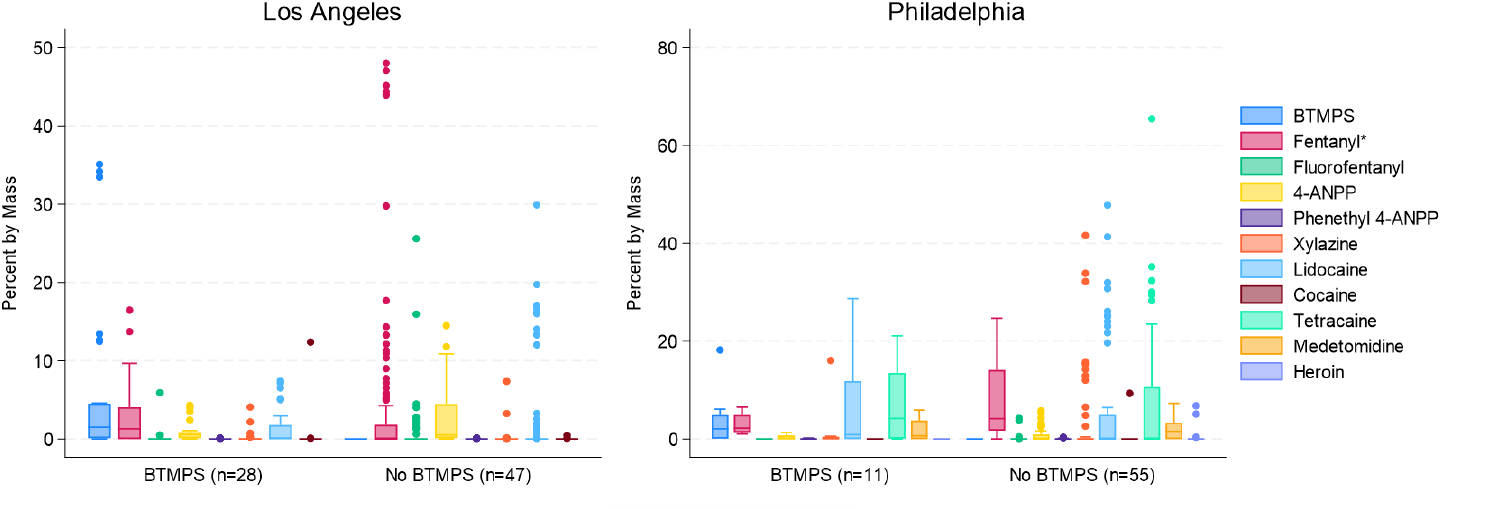
Difference in quantitative results by BTMPS presence for samples from Los Angeles (left) and Philadelphia (right), *p<0.05.

### Participant reports

At community-based drug checking programs in Los Angeles and Philadelphia, participants are invited to complete an optional brief survey when getting a sample tested. Participant surveys were available for eight samples in Los Angeles and four in Philadelphia that contained BTMPS, providing additional qualitative results. Collation of the surveys uncovered some common and troubling themes. One participant stated, “it smelled so bad I could barely smoke it,” while another stated that it made them sick. On subjective assessments of quality, participants rated it from “bunk,” (e.g. very low quality) to moderate (6 on a 10-point scale). One respondent wrote that they believed the sample was “not drugs,” though it did contain 2.3 % fentanyl by mass. Warnings that a pungent odor is correlated to a new fentanyl additive, now known to be BTMPS, have appeared in advisories written by various syringe service and drug checking programs. The smell is alternately described as “fishy,” “like bug spray” or “Raid,” “rubbery,” or “like plastic.”

### Trace residue testing

In addition to the two sites that tested actual drug product, six other sites that used the trace residue collection approach also identified BTMPS through qualitative testing. In July 2024 and August 2024, BTMPS was identified in samples from Delaware, Maryland, Nevada, Washington, and two other cities within California. A total of 23 additional samples (ranging from one to nine per site) were identified. Over half (n=13, 57 %) contained one or more of the local anesthetics discussed above. Six of the samples (26 %) also contained xylazine and three (13 %) contained medetomidine. Other commonly co-detected compounds included fentanyl (n=19, 83 %), 4-ANPP (n=8, 35 %), and acetaminophen (n=11, 48 %). It is important to note that many of these samples were collected from used drug paraphernalia and co-use or cross-contamination of the paraphernalia cannot be ruled out.

## Discussion

The emergence of BTMPS is the most sudden, by way of prevalence, change to the illicit drug supply that we have observed in recent history. Unlike fentanyl and xylazine, it is not a compound that has been sold or understood as a drug with psychoactive properties. And yet within two months of detection in drug checking programs, BTMPS has been identified in at least eight jurisdictions. Where quantitative information is available, the amount of BTMPS in the sample has been shown to exceed the amount of fentanyl, sometimes by a factor of 10 or 20. Knowledge about the specific health risks associated with smoking or injecting BTMPS is limited by its recency in the illicit drug supply, but lethality and serious health risks have been established in occupational health and animal toxicology studies.^6, 7, 11, 12^ Thus, in addition to the well-known health and safety consequences associated with illicit fentanyl use, the sudden introduction of BTMPS raises substantial public health concerns.

Inhalation of BTMPS has been shown to have serious health risks.^6, 11, 12^ This is especially troubling because inhalation is the primary route of administration for fentanyl in Los Angeles (and the West Coast generally).^13-16^ Published lethal dose (LD_50_) values via inhalation range from 0.5 mg/L to 1 mg/L, based on studies in rats.^11, 12, 17^ A report by Australia’s National Industrial Chemicals Notification and Assessment Scheme further notes that rats subjected to inhalation toxicology studies of BTMPS exhibited a range of ailments, including dyspnea, trismus, tremor, and sedation.^12^ BTMPS was found to cause serious, sometimes irreversible, eye damage in rabbits.^11, 12^ People who smoke fentanyl, due to the basic logistics of the act, very often get smoke in their eyes, which smokers report causes a burning or stinging sensation, though in the context of them inhaling the rest of the smoke is perceived as a secondary concern.

In our study, as much as 35 % by mass of what individuals purchased as “fentanyl” was actually BTMPS, a cause for concern if the relationship between exposure and adverse events is dose dependent. With such a sudden and sustained prevalence in the drug supply, users are at risk of repeated, ongoing exposures, which may compound health effects. The sustained prevalence of BTMPS over time can be extrapolated to mean that a substantial portion of people who use fentanyl have been exposed to varying quantities of BTMPS, likely repeatedly.

Although it is not certain why BTMPS is present in recent fentanyl samples, given the high percentages of BTMPS detected in samples, it is possible that illicit drug manufacturers are adding BTMPS to one or more of the fentanyl precursors, at some point in the synthesis process, or to the final product. Its function is unknown as fentanyl was widely synthesized and distributed without BTMPS until very recently, and fentanyl samples without BTMPS continue to be found, indicating uneven use across manufacturers or manufacturing instances. Given the widespread geography of its detection, this addition may be happening at a high level in the supply chain. Also concerning is the fact that many sales listings for BTMPS on surface web e-commerce platforms posted within the past year are qualitatively similar to how some chemical manufacturers have marketed to illicit drug manufacturers, with claims of being able to clear customs, promising high purity and secrecy, etc. Further investigation is warranted and such efforts are currently underway.

It is plausible that BTMPS could be introduced to stabilize fentanyl, or fentanyl precursors, from degradation that could occur from exposure to light or heat during manufacturing, storage, or transportation, and thus increase production yield. For example, 4-ANPP is susceptible to thermal and oxidative decomposition due to the presence of two basic (and, therefore, reactive) nitrogen atoms.^18, 19^ Our quantitative results indicate it is regularly present at amounts that exceed what would be expected from surface contamination, *e.g*., from plastics used in manufacturing.^20^ The relevance of a high prevalence of cocaine and chemically similar local anesthetics is unknown. It is important to note that lidocaine has long been present in certain drug markets.^21, 22^

Despite the substantial toxicity profile, BTMPS has been studied at low doses in rats for a potential role in treating opioid use disorder and other substance use disorders.^3-5^ Prior work has theorized that, given BTMPS is a nicotine antagonist., it could delay onset of opioid withdrawal symptoms, though translational research to humans has not been published.^3, 4^ We hypothesize that the pharmacologic effects demonstrated in animal studies are incidental, rather than a main reason it has been added to illicit fentanyl.^3-5^ As samples with high amounts of BTMPS were described as “bunk,” it does not appear to be a psychotropic drug.

Broadly, drug users are perceiving the sudden onset of BTMPS as highly undesirable and something to be avoided. The fact that quantitated fentanyl samples containing BTMPS had less than half the purity of samples not containing BTMPS corroborates this concern in a setting where drug users prefer higher purity fentanyl with fewer adulterants. But at a prevalence of 41 % in the most recently available data in Los Angeles and nearly 25 % in Philadelphia, for people dependent on fentanyl it is extremely challenging to avoid entirely. Without an option for rapid drug checking (*i.e*., there is no BTMPS test strip), this effort is even more difficult. BTMPS is detectable on Fourier transform infrared spectrometer, which means that community-based programs using this common type of portable spectrometer will generally be able to identify it at point-of-service for clients who present for in-person drug checking. Given the substantial toxicity noted in animal studies and chemical safety datasheets, further research on health effects in humans is urgently needed. Developing and validating clinical tools to identify exposure to BTMPS may also be warranted, to aid clinicians in diagnosing and treating whatever the presentation of BTMPS exposure may be. Due to a lack of testing for BTMPS in vitro or postmortem, should exposure have already caused severe ill effects resulting in morbidity or mortality in someone who involuntarily consumed BTMPS, clinicians would have no way of knowing. Without broad awareness of this new additive, emergency departments and forensic toxicologists may not know to suspect it as a cause. Expanded in vitro and postmortem testing is urgently needed.

The identification of emerging and unusual substances of concern is limited by the number of drug checking programs, an expansion of these programs would facilitate improved detection capabilities. Drug checking programs detected BTMPS before other traditional illicit drug supply data sources, hopefully hastening the time to education and interventions that can protect human health and life. As it is not a controlled substance or something meant for human consumption, in addition to not being part of any standard toxicology panels, BTMPS would not be the focus of testing in criminal investigations. While our study reports results from eight jurisdictions, the spread of BTMPS has been documented in more locations by community-based drug checking programs. According to publicly posted data, between April 10, 2024 and August 26, 2024, the UNC Street Drug Analysis Lab had detected 61 samples that contained BTMPS from California, Colorado, Michigan, New Mexico, New York, North Carolina, Oregon, Washington, and Wisconsin.^23^ As drug checking programs are not widely available, BTMPS is likely present in many more areas.

### Limitations

Community-based drug checking relies on convenience sampling (*i.e*., testing samples that are voluntarily provided) and as such may not represent the wider illicit drug supply in cities, much less regions that do not have drug checking programs. Both the Los Angeles and Philadelphia drug checking programs operate in areas with high volumes of open-air drug sales and use.

Laws on where drug checking is explicitly allowed often permit it only in the context of syringe service programs, which serve only a subset of people who use drugs.^24^ Because drug checking is not universally available, it is possible that BTMPS was present in the US drug supply earlier and simply not detected. Most of the substances tested in this analysis were powder products, rather than pills. As such, more testing of pills is warranted to establish the degree to which BTMPS adulteration is present in markets other than powder fentanyl.

Large molecules – such as sugars and other common bulking agents – were generally neither quantifiable nor detectable in qualitative DART-MS analyses and therefore not reported. Quantitation results include only compounds on the panel, and non-quantified components of a sample can be understood to include bulking agents as well as other active ingredients that are not part of the quantitation panel. Therefore, fentanyl purity on its own may not fully account for a given sample’s potency. The impact of this on the present BTMPS results is likely to be minimal, as *para-*fluorofentanyl (which was quantified) was the only other active fentanyl analog detected in samples with BTMPS. Two of the non-BTMPS samples in Los Angeles included methylfentanyl, so it is possible that the average potency (based on fentanyl and fentanyl analogs) of non-BTMPS samples would be even higher. Additionally, other ingredients with toxicity profiles – for example, aniline that can damage hemoglobin and tramadol can cause nephrotoxicity – were not quantified.^25, 26^ Therefore, beyond the lack of information about BTMPS human pharmacology, it is not possible based on the results of this study to rule out other additives as contributory to adverse effects reported by participants. Yet, the variable and often high prevalence of BTMPS in fentanyl samples is concerning and highlights the need for further studies to establish its effects on human health.

## Conclusions

Prior to April 2024, there was no record of BTMPS in the illicit drug supply. Within four months, it was detected in significant and consistent amounts across the United States. Given the substantial toxicity profile, especially by inhalation, the rapid and broad emergence of this adulterant necessitates further study to characterize and mitigate risks to human health.

## Data Availability

All data produced in the present study are available upon reasonable request to the authors

## Disclaimer

Certain commercial products are identified in order to adequately specify the procedure; this does not imply endorsement or recommendation by NIST, nor does it imply that such products are necessarily the best available for the purpose.

## Funding

A portion of this work was funded by the Division of Overdose Prevention, National Center for Injury Prevention and Control (NCIPC), Centers for Disease Control and Prevention. Authors were also funded by grants from the National Institutes of Health.

## Supplemental Information

### Sample preparation for quantitative testing

Samples for quantitative testing were received as solutions of a few milligrams of drugs in acetonitrile. Upon receipt, qualitative testing by DART-MS was completed after which the solvent was allowed to evaporate off. Once the solvent was evaporated, the weight of drug product was obtained and the sample was reconstituted in methanol containing internal standards (cocaine-d_3_, fentanyl-d_5_, methamphetamine-d_5_, and xylazine-d_6_). The sample was then diluted 1,000-fold in methanol containing internal standard before being analyzed by LC-MS/MS.

A Thermo UltiMate 3000 LC system coupled to a Sciex QTrap4000 or Thermo TSQ Quantis Plus mass spectrometer was used. LC parameters included use of a Biphenyl 2.7 μm x 4.6 mm x 150 mm column (Restek), and a 5 μL injection value. The mobile consisted of methanol with % formic acid (A) and water with 0.1 % formic acid (B). The mobile phase program was i) a 12 min gradient from 95 % B / 5 % A to 100 % A, ii) a 3 min isocratic period at 100 % A, and iii) a 3 min isocratic period at 95 % B / 5 % A. A flow rate of 0.5 mL/min and a column oven temperature of 50 °C was used. In total, the run time for the method was 18 min. Global mass spectrometer parameters included operation in positive ionization with an electrospray needle voltage of 5500 V (Sciex) or 4500 V (Thermo) and a source temperature of 550 °C (Sciex) or 350 °C (Thermo). The mass spectrometers were operated in multiple reaction monitoring (MRM) mode, using the transitions provided below. For each compound a quantitative and qualitative transition were monitored and the peak area of the quantitative transition compared to that of the appropriate internal standard. All ratios were compared to those from a gravimetrically prepared calibration curve in the range of 0.01 μg/mL to 10 μg/mL. Percent by mass values were obtained by correcting the calculated concentration value for the dilution and initial mass of drug product.

**Supplemental Table 1.** MRM transitions, and corresponding internal standards, used for quantitative analysis.

| Compound | Precursor (Da) | Product (Da) | Retention Time (min) | Quant / Qual Transition | Internal Standard | Collision Energy (V) |
| --- | --- | --- | --- | --- | --- | --- |
| 4-ANPP | 281 | 188 | 6.1 | Quant | Fentanyl-d<sub>5</sub> | 40 |
|  | 281 | 105 | 6.1 | Qual | NA | 53 |
| BTMPS | 481.5 | 342.7 | 6 | Quant | Fentanyl-d<sub>5</sub> | 40 |
|  | 481.5 | 140.5 | 6 | Qual | NA | 40 |
| Cocaine | 304.2 | 182 | 5.5 | Quant | Cocaine-d<sub>3</sub> | 40 |
|  | 304.2 | 105 | 5.5 | Qual | NA | 55 |
| Fentanyl | 337 | 188 | 6.4 | Quant | Fentanyl-d<sub>5</sub> | 35 |
|  | 337 | 105.1 | 6.4 | Qual | NA | 50 |
| *para*-Fluorofentanyl | 355 | 188 | 6.3 | Quant | Fentanyl-d<sub>5</sub> | 33 |
|  | 355 | 105 | 6.3 | Qual | NA | 53 |
| Heroin | 370.2 | 328 | 5.3 | Quant | Fentanyl-d<sub>5</sub> | 37 |
|  | 370.2 | 268 | 5.3 | Qual | NA | 40 |
| Lidocaine | 235 | 86 | 4.1 | Quant | Cocaine-d<sub>3</sub> | 33 |
|  | 235 | 58 | 4.1 | Qual | NA | 45 |
| Medetomidine | 201 | 95 | 5.7 | Quant | Xylazine-d<sub>6</sub> | 25 |
|  | 201 | 68 | 5.7 | Qual | NA | 55 |
| Phenethyl 4-ANPP | 385 | 134 | 7.8 | Quant | Fentanyl-d<sub>5</sub> | 50 |
|  | 385 | 105 | 7.8 | Qual | NA | 75 |
| Tetracaine | 265 | 176 | 6 | Quant | Cocaine-d<sub>3</sub> | 30 |
|  | 265 | 120 | 6 | Qual | NA | 50 |
| Xylazine | 221 | 90 | 5.2 | Quant | Xylazine-d<sub>6</sub> | 33 |
|  | 221 | 164 | 5.2 | Qual | NA | 38 |
| Cocaine-d<sub>3</sub> | 307.2 | 105 | 5.5 | Quant | NA | 55 |
| Fentanyl-d<sub>5</sub> | 342 | 188 | 6.4 | Quant | NA | 35 |
| Methamphetamine-d<sub>5</sub> | 155.3 | 96.1 | 3.6 | Quant | NA | 32 |
| Xylazine-d<sub>6</sub> | 227 | 170 | 5.2 | Quant | NA | 50 |

## Notes

### Competing Interest Statement

The authors have declared no competing interest.

### Author Declarations

UCLA IRB determined that the activities described in this analysis, funded by CDC OD2A:LOCAL were public health surveillance and did not constitute human subjects research. The research activities in Philadelphia were approved by the WIRB-Copernicus Group (WCG IRB).

